# Reactive Compensation for Proactive Deficits in Parkinson’s Disease

**DOI:** 10.64898/2026.09.10.26362782

**Authors:** Julia Ficke, Julius Kricheldorff, Stefan Debener, Kathrin Janitzky, Karsten Witt

## Abstract

The dual mechanisms of control (DMC) framework distinguishes between proactive (anticipatory) and reactive (post-conflict) adaptive cognitive control mechanisms. Evidence for proactive deficits in Parkinson’s disease (PD) is inconsistent, and the neurophysiological underpinnings remain poorly understood. We investigated adaptive control in 30 PD patients and 30 demographically similar healthy controls. Adaptive cognitive control was tested using a numerical Stroop test that included manipulations sensitive to proactive and reactive control. In both behavioral and EEG analyses, inducer items that trigger control adaptation were distinguished from diagnostic items, allowing adaptation to be assessed independently of learning effects. Event-related potentials (ERPs) derived from electroencephalography (EEG) captured signatures of proactive (N450) and reactive (Late Positive Complex, LPC) control.

The HC group showed robust proactive control with faster responses compared to reaction times to reactive control condition, whereas PD revealed reduced proactive control, particularly for diagnostic items. ERP analysis indicated a clear N450 in the HC group, whereas the N450 was absent in the PD group. Both groups displayed LPC activity during reactive control, but PD also showed LPC activity in the adaptive proactive control condition, suggesting compensatory reliance on reactive mechanisms. Both, behavioural and ERP analyses provide insights into the dissociation of reactive and proactive control processes in PD. Our findings indicate selective impairments in proactive control, along with preserved reactive control, in patients with PD. Moreover, the presence of the LPC during proactive control trials suggests that the recruitment of reactive control mechanisms may compensate deficits in proactive control in PD.

## Introduction

Parkinson’s disease (PD) is a neurodegenerative disorder that leads to degeneration of dopaminergic neurons, resulting mainly in severe motor symptoms. Cognitive impairments are also frequently part of the symptom spectrum, including impaired cognitive control.^1,2^ Previous work has shown that PD is associated with impaired adaptive control (also referred to as control learning), that is, the dynamic adjustment of cognitive control to changing environmental demands.^3–5^ In the following, we use the term adaptive control consistently. Bravers^6^ dual mechanisms of cognitive control (DMC) framework distinguishes between proactive and reactive control processes. Proactive control is an anticipatory mechanism that enables the resolution of potential information conflicts before they arise. In contrast, reactive control is recruited after conflict has been detected.^7^

Proactive control is commonly conceptualized as the sustained, anticipatory maintenance of goal-relevant information, primarily supported by frontostriatal circuits, which are known to be disrupted in Parkinson’s disease. In contrast, reactive control reflects a transient, stimulus-driven recruitment of control processes, associated with lateral prefrontal and anterior cingulate regions.^6,8^

Conflict tasks such as the Stroop task are commonly used to investigate adaptive control. The Stroop paradigm includes trials that are either congruent (aligned) or incongruent (conflicting), demanding cognitive control to inhibit automatic response tendencies. A specific variant is the numerical Stroop task, in which participants compare two digits that can vary in numerical magnitude and in physical size. Participants are instructed either to indicate the side on which the numerically larger digit appears (numerical task), ignoring physical size, or to indicate the side of the physically larger digit (physical task), disregarding numerical value.^9^ A congruent condition occurs when the numerically larger digit is also physically larger, whereas an incongruent stimulus occurs when the numerically smaller digit appears physically larger. Behavioral response times tend to be faster for congruent stimuli (facilitation) and slower for incongruent stimuli (interference).^10^ Previous research has demonstrated large congruency effects in the numerical Stroop task.^9^

By manipulating the proportion of congruent trials, adaptive control can be examined.^3,11,12^ The list-wide proportion congruency effect (LWPC) reflects global proactive control, while the item-specific proportion congruency effect (ISPC) assesses reactive control.^7,13^ Importantly, adaptive control can be disentangled from lower-level learning processes using diagnostic items not directly manipulated in congruency frequency.^3^ This inducer-diagnostic design has been recommended by Braem et al.^3^. Inducer items are presented with biased frequencies to trigger adaptive control, while diagnostic items appear with equal frequency and are free from stimulus–response or frequency-based associations. This manipulation allows for a valid assessment of adaptive control, minimizing confounding learning effects.^14^ Analyses of inducer items are therefore also reported to verify the effectiveness of the manipulation and to aid interpretation of the diagnostic item effects.

Studies investigating proactive control in PD have yielded inconclusive results. While some studies reported impairments,^11,12^ others failed to report similar effects.^15,16^ This variability may arise from differences in task demands, such as the extent to which proactive control is required, as well as from patient-related factors, including disease severity, dopaminergic medication status, and individual differences in cognitive resources. In particular, dopaminergic medication has been shown to differentially affect cognitive control processes depending on task demands and baseline dopamine levels, consistent with the dopamine overdose hypothesis.^8,17^ Moreover, proactive control is thought to rely on sustained goal maintenance processes supported by frontostriatal circuitry,^6^ which may be differentially compromised across patients and experimental contexts.

Bonnin et al.^18^ found no LWPC effect in medicated PD participants. The LWPC effect refers to a reduction of Stroop interference in high-versus low-conflict contexts and reflects intact proactive control. The absence of sustained proactive control in these participants may be due to limitations in attentional resources, which are heavily taxed during proactive control. Ruitenberg^19^ reported that PD primarily affects proactive control in the Eriksen flanker task, with reactive control being less impaired. In contrast, Rodriguez-Raecke et al.^16^ found no impairment of the congruency sequence effect in patients with PD, using the Eriksen flanker task.

Although behavioural studies have examined adaptive control in PD,^5,18,20^ investigations into its neurophysiological correlates remain scarce. ERPs, for example, offer high temporal resolution that allows proactive and reactive control processes to be dissociated, as proposed by the DMC model. In Stroop tasks, proactive control has been linked to the N450, a frontocentral negative component occurring 350-500ms after stimulus onset, which has been associated with conflict monitoring and the detection of response conflict.^10,11,21^ The N450 originates in the anterior cingulate cortex (ACC).^22^ While direct evidence on N450 alterations in Parkinson’s disease (PD) is limited, studies on related conflict-monitoring components, such as the error-related negativity (ERN), have shown reduced amplitudes in PD, suggesting impairments in medial frontal and frontostriatal monitoring mechanisms.^23^

Reactive control is associated with a late positive component (LPC) over centroparietal regions (600-900ms). This component overlaps functionally with the P3b and is thought to reflect processes related to attentional allocation, context updating, and response selection.^21,24,25^ Importantly, findings regarding late positive ERP components in PD are heterogeneous. While some studies have reported reduced or delayed P3 amplitudes, others have found relatively preserved LPC/P3b activity depending on task demands and patient characteristics.^25,26^ This variability suggests that reactive control processes may be differentially affected in PD, potentially depending on task demands and patient characteristics.

In a recent study^4^, we provided evidence that proactive control processes are impaired in patients with PD. Specifically, we found that patients with PD exhibited reduced preparatory activity associated with proactive control, as indicated by EEG time-frequency analysis. The results complement previous findings suggesting that PD patients exhibit difficulties with anticipatory adjustments in cognitive control,^5,18,19,27^ potentially reflecting dopaminergic dysfunction within a network that includes the anterior cingulate cortex (ACC), a key region for proactive control.^22^

The present study builds on prior ERP research by tracking impairments in proactive and reactive control with precise temporal resolution. We hypothesized that the impaired proactive control observed in patients with PD is associated with an altered N450 component, whereas preserved reactive control is reflected by an intact LPC component. In addition, we conducted a frequentist reanalysis of behavioural data to validate previous Bayesian findings.^4^ Together, these approaches allow for a more precise characterization of proactive control deficits in PD and reveal new aspects of the neural mechanisms underlying cognitive control impairments.

## Methods

This prospective, monocentric study was conducted at the University Department of Neurology outpatient clinic at the Evangelisches Krankenhaus Oldenburg, Germany. The study protocol was reviewed and approved by the Medical Ethics Committee of Carl von Ossietzky University Oldenburg (file number 2020-133) and preregistered in the German Register for Clinical Studies (DRKS00023020). The data were collected between December 2020 and December 2021.

### Study Participants

This study included 30 participants with PD diagnosed according to MDS clinical diagnostic criteria and 30 healthy control participants who were demographically similar in age and sex (50–75 years).^28^ Four healthy control participants were initially excluded from the analysis because they did not follow instructions to respond using only their right hand. These exclusions were identified after initial data collection, and four replacement control participants were subsequently recruited and included into the final analysis to maintain the planned sample size (Table 1). Additionally, one PD participant was excluded for not being attentive throughout the task. All participants were recruited according to the following criteria: absence of neurological or psychiatric disease (other than PD), absence of dementia (Mini-Mental State Exam > 26 points), right-handedness, fluency in German and normal or corrected-to-normal vision. All participants with PD were tested on their standard dopaminergic medication regimen, but anticholinergic medication was an exclusion criterion. The Movement Disorders Society Unified Parkinson Disease Rating Scale (MDS-UPDRS) Part III was used to assess the severity of motor symptoms in PD patients. All participants gave verbal and written informed consent before the experiment, in agreement with the Declaration of Helsinki.^29^

**Table 1:** Descriptive data for demographic and clinical characteristics of participants by group. MMSE = Mini-Mental State Examination (0–30; higher scores indicate better cognition). MDS-UPDRS-III = Movement Disorders Society Unified Parkinson’s Disease Rating Scale–Motor Section (0–132; higher scores indicate greater motor impairment). Educational years were calculated as the total number of school years plus years of non-academic training or higher education.

| | Parkinson Group<br>Count/ Mean ± SD (range) | Healthy Group<br>Count/ Mean ± SD (range) |
| --- | --- | --- |
| Number | 29 | 30 |
| Age (years) | 64.0 ± 9.6 (46 -80) | 59.4 ± 6.8 (50-74) |
| Education Years | 16.2 ± 3.0 (12 -22) | 16.0 ± 3.2 (11-22) |
| Sex (m/w) | 21/ 8 | 18/ 12 |
| MMSE | 29.2 ± 1.1 (26-30) | 29.1 ± 1.0 (27-30) |
| MDS-UPDRS III (motor score) | 12.0 ± 7.0 (2-37) | n.a. |
| Disease duration (years) | 5.3 ± 3.7 (n.a.) | n.a. |
| Hoehn and Yahr Stage | 2 ± 0.5 (1-3) | n.a. |

PD individuals were significantly older than those in the healthy control group, t (29) = 2.18, p = 0.036. The groups did not differ in education and gender ratio (see Table 1).

### Task and Stimulus

For adaptive control assessment, we used the numerical Stroop task.^9^ Participants had to choose the numerically larger of two digits while their physical size was manipulated (one digit was displayed larger than the other). For a congruent condition, the relationship between the numerical value and the physical size matched (the numerically smaller number appeared physically smaller) and incongruent conditions were implemented by a mismatch (the numerically smaller number was presented in larger physical size). The experiment was coded in OpenSesame.^30^ For the comparisons, we used the numbers one to nine (excluding five) with numerical distances of one and two (for further details, Supplement Table 1).^9^

The experiment consisted of eight blocks of 134 trials each: four blocks were designed to study the LWPC effect and four blocks were designed to investigate the ISPC effect. Each block consisted of 94 inducer items and 40 diagnostic items (balanced in terms of congruent and incongruent item presentation), resulting in a 70:30 ratio of inducer to diagnostic items.

LWPC refers to a manipulation in which the proportion of congruent versus incongruent trials is defined at the block (i.e., list-wide) level. Accordingly, two blocks were mostly congruent (MC, 80% congruent, 20% incongruent) and two blocks were mostly incongruent (MI, 80% incongruent, 20% congruent). This manipulation creates a global conflict context that is assumed to primarily engage proactive control processes. From here on, the terms MC_LWPC_ and MI_LWPC_ refer to mostly congruent and mostly incongruent LWPC block contexts. For the ISPC effect, the overall number of congruent and incongruent trials in each block was equal (50:50). However, certain number combinations were biased to be mostly congruent or mostly incongruent. Specifically, either the four smaller number combinations (12, 13, 24, 34) or the four larger number combinations (67, 68, 78, 89) were presented as 80% incongruent or 80% congruent, with the opposite ratio applied to the other set of combinations. Two of the four combinations were randomly assigned to the inducer category and two to the diagnostic category, ensuring balanced presentation of diagnostic items. This manipulation is assumed to primarily engage reactive control processes. From here on, the terms MC_ISPC_ and MC_ISPC_ refer to the mostly congruent and mostly incongruent ISPC item categories, respectively.

Inducer items establish the congruency context (i.e., the manipulation that drives learning of the contingency structure), whereas diagnostic items are used to measure the resulting control adjustments without contributing directly to the learned contingencies. Stimulus presentations within a block were pseudorandomized (see Supplementary Information 1) to ensure that diagnostic items were presented regularly across the block and to assess the effects of the inducer manipulation.

### Procedure

The experiment began with a practice block containing 30 stimuli with feedback. To rule out sequence effects in the analysis, the test blocks were randomly presented to the participants in different orders. Participants were given the opportunity to take a break between each block.

A stimulus consisted of two digits displayed simultaneously in the center of a 27-inch screen. The stimuli were black on a white background. A trial began with a fixation point displayed for 300 to 600 ms, followed for a maximum of 2000 ms by two digits to the left and right of the fixation point. Participants were asked to indicate on which side the numerically larger number appeared by pressing the left arrow key for the left number and the right arrow key for the right number on a standard computer keyboard using the index and the middle finger of the right hand. After a response (or if more than 2000 ms passed), the number combination disappeared and was followed by a white screen for 800 ms until the next stimulus (visualized in Figure 1).

**Figure 1.**
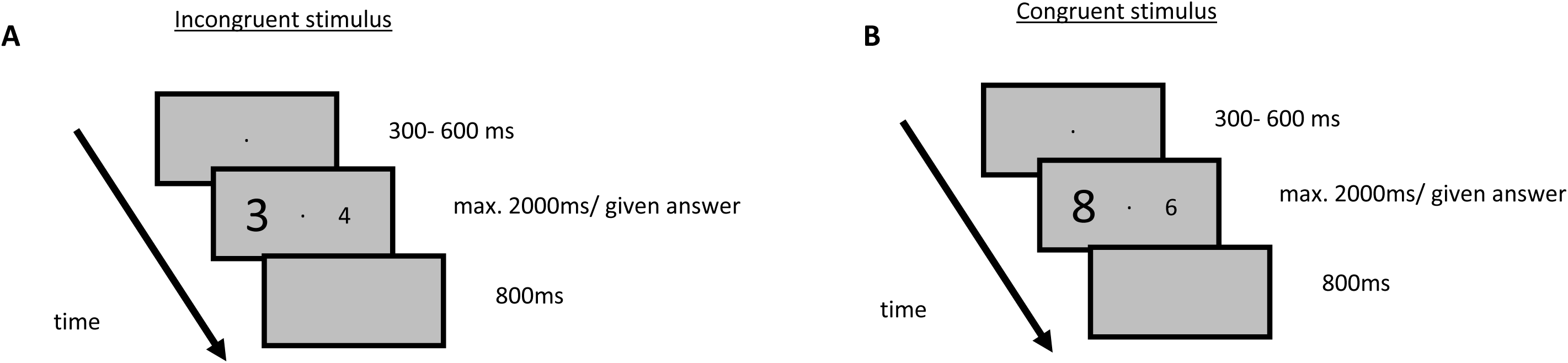
Trial sequence for congruent and incongruent stimuli. In incongruent stimuli. (Panel A), the numerical size of the digit does not match its physical size, inducing conflict; in congruent stimuli (Panel B), numerical and physical size correspond. Participants responded using the left and right arrow keys, indicating which digit was numerically larger, with the correct response corresponding to the side of the numerically larger digit (e.g., right arrow for an incongruent trial and left arrow for a congruent trial in the illustrated examples). Each trial included a fixation (300–600 ms), stimulus presentation (max. 2000 ms), and an inter-trial interval (800 ms).

### Electrophysiological recording

The EEG was recorded continuously (actiChamp plus by Brain Vision) with a sampling rate of 1000 Hz from 128 active Ag/AgCl electrodes with the FCz electrode as the reference. Scalp impedances were kept below 10 kΩ.

### Statistical Analysis

Behavioural data were analysed using R (Version 2026.01.0+392^31^). Behavioural data were merged with a participant database including demographic information and, for Parkinson’s patients, Unified Parkinson’s Disease Rating Scale (UPDRS) scores.^32^ Preprocessing steps were conducted to ensure data quality. Response times below 200 ms were removed to exclude anticipatory responses, resulting in the removal of 20 trials (0.03% of the data). Reaction times were log-transformed prior to analysis to improve normality and model assumptions.

Linear mixed-effects models (LMMs) were fitted using the lmerTest package.^33^ All models included by-subject and by-item random intercepts to account for repeated-measures dependencies in the data.

Fixed-effects structures were defined a priori using orthogonal contrast coding implemented via the hypr package.^34^ This approach enabled direct hypothesis-driven tests of (i) congruency effects, (ii) context-dependent adaptive control within the list-wide (MC_LWPC_ and MI_LWPC_) and item-specific (MC_ISPC_ and MI_ISPC_) proportion congruency manipulations, and (iii) their interactions with group (healthy controls vs. Parkinson’s disease). Separate but structurally aligned contrast schemes were defined for list-wide and item-specific conditions to allow independent estimation of control-related effects across task contexts. Here, we calculated the ‘Main Effect of Congruency’ as the difference between congruent and incongruent items across the two conflict conditions (MC and MI). ‘Main Effect of ISPC/LWPC Manipulation’ was operationalized as the overall difference in RT between items of the MI and MC conflict contexts. Finally, we operationalized adaptive control as the difference in Congruency effects (congruent–incongruent) between the MC and MI LWPC block contexts or ISPC item categories – termed ‘Congruency x ISPC/LWPC Interaction’. For a complete list of contrast definitions, see Supplement Table 2. The analyses were performed for each group (healthy controls vs. Parkinson’s disease) and item (diagnostic vs. inducer) group separately.

In addition to a full factorial model testing interactions between group, congruency, and condition, separate LMMs were estimated for inducer and diagnostic conditions as well as for list-wide and item-specific datasets. This model decomposition allowed us to assess whether the adaptive control effects generalized across task contexts and trial types. Statistical inference was based on t-statistics and p-values derived from Satterthwaite’s approximation of degrees of freedom as implemented in lmerTest. Wald-type confidence intervals were computed for all fixed effects. No correction for multiple comparisons was applied, as all tests were derived from a unified, pre-specified contrast framework. Error rates were analysed separately to evaluate potential speed–accuracy trade-offs. Data visualisation was performed using the ggplot2 package.^35^

To address potential influences of disease severity and cognitive status on cognitive control effects, additional exploratory covariance analyses were conducted within the Parkinson’s disease group. Subject-specific estimates of cognitive control effects were extracted from the mixed-effects models and regressed on clinical measures (UPDRS), global cognitive status (MMSE), and years of education using linear regression models.

## ERP Analysis

### Pre-processing

EEG data were analysed in MATLAB using the EEGLAB toolbox.^36^ EEG data were resampled from 1000 to 250 Hz, and high-pass filtered at 0.1 Hz. Noisy channels were removed by the clean_rawdata function^37^ with the following criteria: FlatlineCriterion = 5 (channels flatlining for ≥5 s were removed), ChannelCriterion = 0.8 (channels with >80% correlation to others were rejected), and LineNoiseCriterion = 5 (channels with excessive 50 Hz line noise were removed). Removed channels were subsequently interpolated using spherical spline interpolation to restore the full montage. Artifact Subspace Reconstruction (ASR) was applied to detect and remove transient high-amplitude noise segments. ASR operates by identifying bursts of abnormal activity relative to clean reference data (here, the rest of the recording) and reconstructing these segments from the clean subspace. After ASR, independent component analysis (ICA) was performed to decompose the EEG into independent sources, allowing the identification and removal of residual blink- and muscle-related components. To improve the topographical selectivity of the data, a Laplacian was performed using the CSD Toolbox.^38^ This method emphasizes local electrical activity at each electrode while attenuating activity spread from distant sources, making topographical patterns of EEG signals more precise. A 30Hz low-pass filter was used to eliminate high-frequency noise. After preprocessing, the cleaned continuous EEG data were segmented into epochs from 200 ms pre-stimulus to 1500 ms post-stimulus for correctly answered trials, with baseline correction applied using the mean of the 200 ms period prior to stimulus onset.

After preprocessing, one participant in the PD group was excluded due to insufficient data quality, characterized by extended periods of non-task-related activity, and one participant in the healthy control (HC) group due to technical problems during part of the recording. This left a final sample of 28 PD participants and 29 HC participants for the LWPC and ISPC analyses.

### Stimulus-locked analysis

For the ERP analysis, we focused on the N450 and the late positive complex (LPC) components. The latency window for these two components was determined using the collapsed localizer approach (Supplementary Information 2).^39,40^ The amplitude of the N450 was measured as the mean voltage in the epoch from 352–500 ms, and the amplitude of the LPC as the mean voltage between 560 and 944 ms.

Difference waves were computed for each condition (MI and MC in LWPC and ISPC blocks) and group (HC and PD) by subtracting the ERP waveform for congruent stimuli from that for incongruent stimuli. This allowed us to examine how cognitive control processes differ across conditions and groups. Electrodes selected for this analysis varied by component, with the N450 being analysed using electrodes FCz, FC1, CP1, CP2, Cz, FC2, C1, CPz, C2, whereas the LPC was examined at POO1, POO2, P1, P2, POz, Pz, CPz, PPO1h, PPO2h, CPP1h, and CPP2h. Electrode selection was defined a priori based on previous literature on conflict processing and cognitive control.^21^

For time-resolved analysis the difference between the LWPC- and ISPC-related Stroop effect curves were compared within each group using two-sided dependent t-tests. P-values were corrected for multiple comparisons using the Benjamini-Hochberg FDR procedure applied within predefined, component-specific time windows.

To assess group differences in condition-related ERP modulation, linear mixed-effects models were computed on mean component amplitudes. These models included group (HC vs. PD) and condition (MC_LWPC_, MI_LWPC_, MC_ISPC_, MI_ISPC_) as fixed factors, as well as their interaction term, allowing direct statistical testing of group differences in condition effects at the level of ERP components.

## Results

### Overall mixed effects model

Descriptive reaction times (means and standard deviations) for all conditions are reported in Supplementary Table 3. Reaction times (log-transformed) were analysed using linear mixed-effects models with pre-specified, hypothesis-driven contrast coding, including fixed effects of group (Parkinson’s disease vs. healthy controls), congruency, and condition, as well as their interactions (see Table 2 and Supplementary Table 4). The overall mixed-effects model simultaneously tested congruency effects and condition-based modulations associated with both the LWPC (MC_LWPC_, MI_LWPC_) and ISPC (MC_ISPC_, MI_ISPC_) manipulations. All reported p-values correspond to pre-specified linear contrasts defined a priori within the mixed-effects model.

**Table 2:** Selected fixed effects from the linear mixed-effects model examining the influence of group (Parkinson’s disease vs. healthy controls), congruency, and condition on log-transformed reaction times. Estimates (b), standard errors (SE), t-values, p-values, and effect sizes (r) are reported. MC_LWPC_ and MI_LWPC_ denote the mostly congruent and mostly incongruent LWPC block contexts, respectively. _MCISPC_ and _MIISPC_ denote the mostly congruent and mostly incongruent ISPC item categories. The _MCISPC_ condition served as the reference level and was therefore not displayed. Random intercepts were included for subjects and items. The complete model output, including all fixed effects, is provided in the Supplementary Materials.

| Effect | b | SE | t | p | r |
| --- | --- | --- | --- | --- | --- |
| Group × Congruency | -0.016 | 0.007 | -2.29 | 0.022 | 0.009 |
| Group × Condition ($MI_{ISPC}$) | 0.021 | 0.007 | 3.01 | 0.003 | 0.012 |
| Group × Condition (MC) | -0.011 | 0.005 | -1.99 | 0.046 | 0.008 |
| Group × Condition (MI) | -0.014 | 0.007 | -2.10 | 0.035 | 0.008 |
| Congruency × Condition ($MI_{ISPC}$) | -0.048 | 0.007 | -7.01 | < .001 | 0.028 |
| Congruency × Condition (MC) | 0.007 | 0.007 | 1.06 | 0.291 | 0.004 |
| Congruency × Condition (MI) | -0.045 | 0.007 | -6.60 | < .001 | 0.026 |
| Group × Congruency × Condition ($MI_{ISPC}$) | 0.012 | 0.010 | 1.28 | 0.199 | 0.005 |
| Group × Congruency × Condition (MC) | -0.009 | 0.010 | -0.92 | 0.356 | 0.004 |
| Group × Congruency × Condition (MI) | 0.005 | 0.010 | 0.52 | 0.606 | 0.002 |

A significant main effect of group indicated overall slower responses in Parkinson’s disease compared to healthy controls (b = 0.106, SE = 0.047, t(58.6) = 2.24, p = 0.029). A robust main effect of congruency confirmed longer response times for incongruent relative to congruent trials (b = 0.110, SE = 0.005, t(64353.1) = 22.62, p < 0.001). Condition further modulated response times, with significant effects for MC (p = 0.014) and MI (p < 0.001), indicating context-dependent modulation of performance.

Significant interactions were observed between group and congruency (p = 0.022) as well as between group and condition (p < 0.05). Congruency further interacted with condition, with strong effects in MI_ISPC_ and MI_LWPC_ conditions (both p < 0.001), but not in MC conditions (p = 0.291). No significant three-way interaction was observed (all p values > 0.19), indicating no evidence for higher-order modulation.

To further characterise adaptive control mechanisms, contrast-based linear mixed-effects models were estimated separately for list-wide and item-specific manipulations and for inducer and diagnostic trials (Table 3).

**Table 3:** Linear mixed-effects model results for list-wide and item-specific Stroop effects in healthy controls (HC) and Parkinson’s disease participants (PD). The table reports fixed effects from four separate mixed-effects models (List-wide Inducer, List-wide Diagnostic, Item-specific Inducer, Item-specific Diagnostic). For each model, effects are shown separately for healthy controls (HC)) and Parkinson’s disease participants (PD). Estimates (b), standard errors (SE), t-values, p-values, and marginal effect sizes (r) are reported. Positive and negative values reflect log-transformed reaction times (Log RT). The main effect of congruency reflects the overall difference between incongruent and congruent trials. Main effects of LWPC and ISPC reflect the influence of the respective context manipulation, whereas Congruency × LWPC and Congruency × ISPC interactions represent the respective proportion congruency effects. List-wide and item-specific effects are derived from orthogonal contrast coding.

| Effect | b | SE | t | p | r |
| --- | --- | --- | --- | --- | --- |
| LW Inducer |  |  |  |  |  |
| Main Effect of Congruency (HC) | -0.097 | 0.004 | -22.92 | < .001 | .152 |
| Main Effect of LWPC Manipulation (HC) | 0.007 | 0.004 | 1.65 | .099 | .011 |
| Congruency x LWPC Interaction (HC) | -0.030 | 0.004 | -7.08 | < .001 | .048 |
| Main Effect of Congruency (PD) | -0.081 | 0.004 | -19.08 | < .001 | .127 |
| Main Effect of LWPC Manipulation (PD) | 0.009 | 0.004 | 2.13 | .033 | .014 |
| Congruency x LWPC Interaction (PD) | -0.024 | 0.004 | -5.71 | < .001 | .038 |
| **LW Diagnostic** |  |  |  |  |  |
| Main Effect of Congruency (HC) | -0.095 | 0.005 | -17.64 | < .001 | .181 |
| Main Effect of LWPC Manipulation (HC) | 0.016 | 0.005 | 3.00 | .003 | .031 |
| Congruency x LWPC Interaction (HC) | -0.020 | 0.005 | -3.82 | < .001 | .040 |
| Main Effect of Congruency (PD) | -0.083 | 0.005 | -15.38 | < .001 | .158 |
| Main Effect of LWPC Manipulation (PD) | -0.007 | 0.005 | -1.34 | .179 | .014 |
| Congruency x LWPC Interaction (PD) | -0.012 | 0.005 | -2.24 | .025 | .023 |
| **IS Inducer** |  |  |  |  |  |
| Main Effect of Congruency (HC) | -0.094 | 0.004 | -22.41 | < .001 | .149 |
| Main Effect of ISPC Manipulation (HC) | 0.025 | 0.004 | 5.97 | < .001 | .040 |
| Congruency x ISPC Interaction (HC) | -0.026 | 0.004 | -6.28 | < .001 | .042 |
| Main Effect of Congruency (PD) | -0.078 | 0.004 | -18.38 | < .001 | .123 |
| Main Effect of ISPC Manipulation (PD) | -0.002 | 0.004 | -0.41 | .678 | .003 |
| Congruency x ISPC Interaction (PD) | -0.022 | 0.004 | -5.09 | < .001 | .034 |
| **IS Diagnostic** |  |  |  |  |  |
| Main Effect of Congruency (HC) | -0.088 | 0.005 | -16.26 | < .001 | .167 |
| Main Effect of ISPC Manipulation (HC) | 0.018 | 0.006 | 3.23 | .001 | .033 |
| Congruency x ISPC Interaction (HC) | -0.009 | 0.005 | -1.72 | .085 | .018 |
| Main Effect of Congruency (PD) | -0.092 | 0.005 | -16.84 | < .001 | .172 |
| Main Effect of ISPC Manipulation (PD) | -0.010 | 0.006 | -1.72 | .086 | .018 |
| Congruency x ISPC Interaction (PD) | -0.009 | 0.005 | -1.66 | .097 | .017 |

### LWPC Task

In inducer trials, robust congruency effects were observed in both healthy controls and Parkinson’s disease participants (HC: b = −0.097, SE = 0.004, t = −22.92, p < 0.001; PD: b = −0.081, SE = 0.004, t = −19.08, p < 0.001). A significant list-wide modulation was present in both groups (HC: b = −0.030, SE = 0.004, t = −7.08, p < 0.001; PD: b = −0.024, SE = 0.004, t = −5.71, p < 0.001), indicating context-dependent adjustment of conflict processing in high-conflict contexts. In diagnostic trials, a significant list-wide proportion congruency (LWPC) effect was observed in healthy controls (b = −0.020, SE = 0.005, t = −3.82, p < 0.001), as well as in Parkinson’s disease participants (b = −0.012, SE = 0.005, t = −2.24, p = 0.025), although the effect was reduced in magnitude in the PD group.

### ISPC Task

In inducer trials, strong item-specific control effects were observed in both groups (HC: b = −0.026, SE = 0.004, t = −6.28, p < 0.001; PD: b = −0.022, SE = 0.004, t = −5.09, p < 0.001), alongside significant item-specific context modulation (HC: b = 0.025, SE = 0.004, t = 5.97, p < 0.001; PD: b = −0.002, SE = 0.004, t = −0.41, p = 0.678).

In diagnostic trials, item-specific effects were weaker and less consistent. While item-specific modulation reached significance in healthy controls (b = 0.018, SE = 0.006, t = 3.23, p = 0.001), no reliable effects were observed in Parkinson’s disease participants (b = −0.010, SE = 0.006, t = −1.72, p = 0.086). The corresponding item-specific proportion congruency (ISPC) effects were not significant in either group (HC: b = −0.009, SE = 0.005, t = −1.72, p = 0.085; PD: b = −0.009, SE = 0.005, t = −1.66, p = 0.097), indicating only weak evidence for item-specific adaptive control in diagnostic trials.

Together, these findings indicate that both groups showed conflict adaptation in inducer items for both LWPC and ISPC manipulations. In diagnostic items, the LWPC effects remained statistically significant in both groups, although the effect was numerically smaller in the PD group. In contrast, ISPC effects in diagnostic items did not reach statistical reliability. An overview of the data is provided in Figure 2.

**Figure 2.**
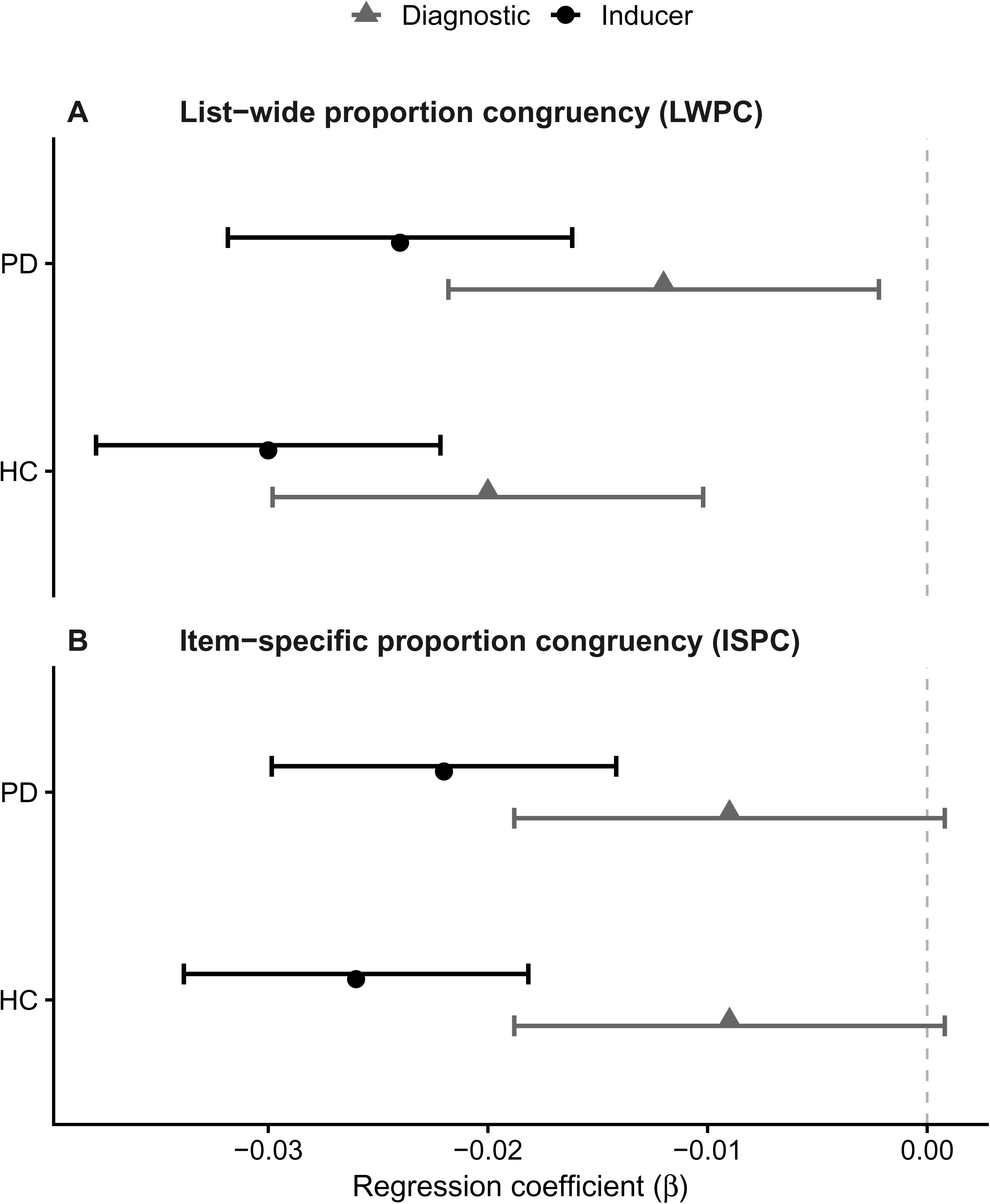
Adaptive control effects estimated from linear mixed-effects models. Regression coefficients (β) and 95% confidence intervals are shown for list-wide proportion congruency (LWPC, panel A) and item-specific proportion congruency (ISPC, panel B) effects in healthy control participants and participants with Parkinson’s disease. Circles indicate inducer-item effects and triangles indicate diagnostic-item effects. Negative coefficients reflect reduced Stroop interference in high-conflict relative to low-conflict contexts.

### Exploratory analyses

Exploratory analyses within the Parkinson’s group examined whether cognitive control effects were associated with UPDRS, MMST, or years of education (Table 4). For list-wide effects, UPDRS showed a small positive association (β = 0.012, p = 0.042), while MMST and education were not significant. No associations were found for item-specific effects (all p > 0.14).

**Table 4:** Regression analyses in the Parkinson’s group relating cognitive control effects to UPDRS, MMST, and education. Effects are based on subject-specific model estimates.

| Model | Predictor | $\beta$ | SE | t | p |
| --- | --- | --- | --- | --- | --- |
| LW Condition | UPDRS | 0.012 | 0.006 | 2.15 | 0.042 |
| LW Condition | MMST | 0.002 | 0.034 | 0.06 | 0.954 |
| LW Condition | Education | 0.001 | 0.011 | 0.10 | 0.923 |
| IS Condition | UPDRS | 0.009 | 0.006 | 1.49 | 0.149 |
| IS Condition | MMST | -0.006 | 0.038 | 0.16 | 0.874 |
| IS Condition | Education | 0.001 | 0.012 | 0.10 | 0.917 |

### Error rates

Error rates were additionally analysed to rule out potential speed–accuracy trade-offs. Overall, error rates were low across conditions. Controls showed minimal error rates in both congruent (M = 0.014, SD = 0.009) and incongruent trials (M = 0.016, SD = 0.014), while Parkinson’s disease patients showed slightly higher error rates in congruent (M = 0.030, SD = 0.024) and incongruent trials (M = 0.032, SD = 0.032). Importantly, no pattern indicative of a speed–accuracy trade-off was observed. These findings are consistent with previous analyses of the same dataset reported by Kricheldorff et al.^4^, who also found no evidence that faster responses were accompanied by increased error rates.

### Event-related brain potentials

In the ERP analysis, we focused on the two components N450 and LPC. The N450 showed a clear frontocentral distribution, peaking around 300–500 ms (see Supplementary Figure 1). Incongruent trials elicited more negative amplitudes than congruent trials, most pronounced at FCz and Cz. This effect was evident in controls and attenuated in patients with PD, indicating reduced conflict-related modulation in the N450 time window. Global ERP scalp topographies (Supplementary Figure 3) further confirmed the frontocentral distribution of the N450 across both groups and conditions.

Regarding the analysis of the N450 for the stimulus-locked proactive control condition, we found a significant N450 (t (28) = 2.62, p = 0.013, d_z = 0.494) in the HC group but not in the PD group (t (28) = -0.43, p = 0.674, d_z = -0.079; Figure 3). Healthy control participants showed the N450 effect, which reflects the typical ERP modulation associated with conflict processing.^10^ PD participants showed no comparable effect. For the N450 component, the linear mixed-effects model revealed a significant main effect of Group (F(1,84) = 21.16, p < 0.001), indicating overall differences in N450 amplitude between healthy controls and Parkinson’s disease participants. The main effect of Condition was not significant (F(1,84) = 1.22, p = 0.273). The Group × Condition interaction did not reach significance (F(1,84) = 2.90, p = 0.092), indicating no robust evidence for group-specific modulation of the conflict effect at the N450 level.

**Figure 3.**
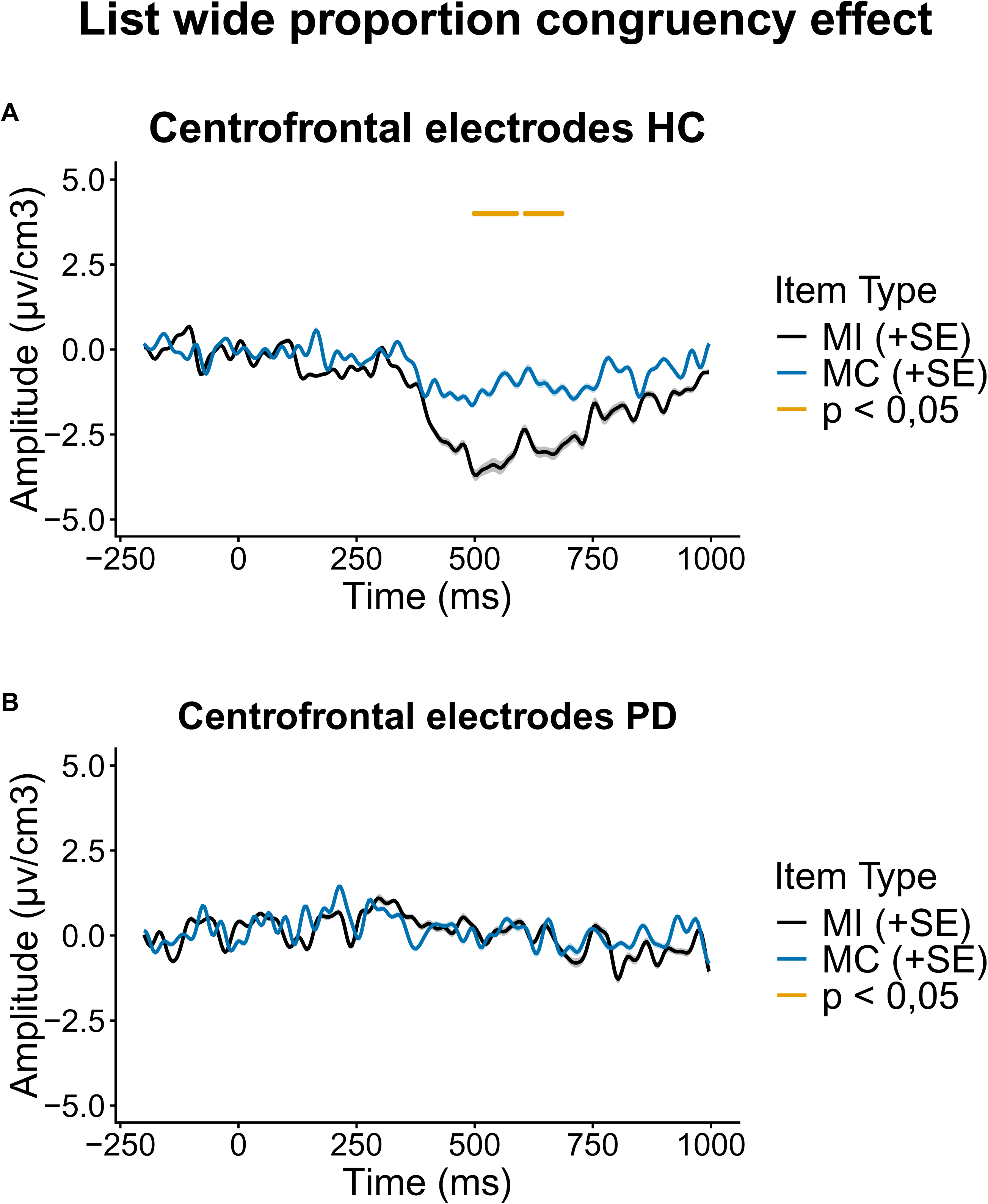
ERP Results for the LWPC effect: Grand-average ERP waveforms at centrofrontal channels for MI_LWPC_ (black) and MC_LWPC_ (blue) conditions in the LWPC task, separately for healthy controls (HC, panel A) and Parkinson’s disease participants (PD, panel B). The x-axis represents time (ms), with 0 ms indicating stimulus onset, and the y-axis shows amplitude (µV/cm³). Shaded areas represent the standard error of the mean (±SE). Time intervals with significant differences between conditions (p < 0.05) are indicated by orange markers. Statistical analyses were conducted using linear mixed-effects models (LMMs) with Group (HC vs. PD) and Condition (MI vs. MC) as fixed effects and random intercepts for participants. Sample size was N = 28 per group. Each waveform represents the grand-average ERP across participants; thus, values reflect mean amplitudes per time point across individuals.

The stimulus-locked ERP waveforms showed a clear LPC over parieto-occipital sites (Supplementary Figure 2), with a typical posterior distribution across both groups. A visual inspection suggests condition-related differences between congruent and incongruent trials in both controls (HC) and Parkinson’s disease (PD) participants, while the overall LPC topography appeared comparable between groups. This posterior distribution is further illustrated by global ERP scalp topographies in Supplementary figure 3.

In the analysis of the LPC in the reactive control condition, we found a significant LPC in the stimulus-locked data in both the HC participants (t (28) = -2.79, p=0.009, d_z=-0.519) and the patients with PD (t (28) = -2.233, p=0.017, d_z=-0.397). Thus, both groups showed an ERP signature of the LPC in the MI item trials, consistent with reactive control processes (Figure 4). For the LPC component, the linear mixed-effects model revealed a significant main effect of Condition (F(1,84) = 9.35, p = 0.003), indicating a reliable modulation of LPC amplitudes by congruency. The main effect of Group was not significant (F(1,84) = 1.81, p = 0.182). The Group × Condition interaction was also not significant (F(1,84) = 0.05, p = 0.825), suggesting that the condition effect was comparable across healthy controls and patients with PD.

**Figure 4.**
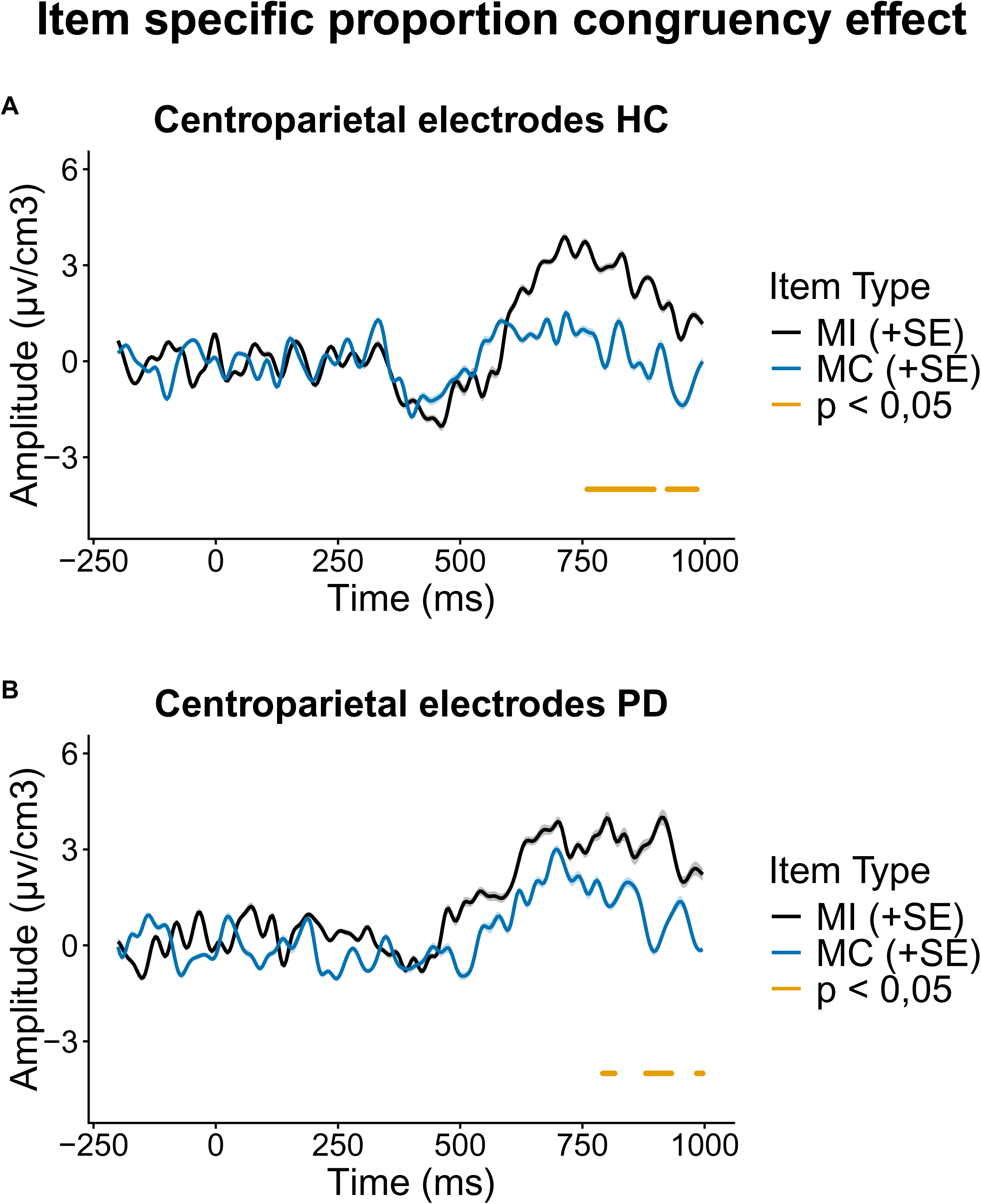
ERP Results for the ISPC effect: Grand average ERP waveforms recorded from centroparietal channels for Healthy Controls (HC, panel A) and Parkinson’s Disease patients (PD, panel B). The x-axis represents time (ms), 0ms indicates the event onset, and the y-axis shows amplitude (uv/cm3). Black and blue lines correspond to different item types (MI_ISPC_ and MC_ISPC_), with shaded areas representing the standard error (SE). Orange horizontal markers indicate time windows with significant differences between conditions (p < 0.05). Statistical analyses were conducted using linear mixed-effects models (LMMs) with Group (HC vs. PD) and Condition (MI vs. MC) as fixed effects and random intercepts for participants. Sample size was N = 28 per group. Each waveform represents the grand-average ERP across participants; thus, values reflect mean amplitudes per time point across individuals.

Despite the absence of a significant N450 effect, patients with PD showed evidence of proactive control adaptation in the behavioral data. In addition, a separate exploratory analysis was conducted to further examine LPC activity in the parietal region of interest within the predefined latency range associated with reactive control (Figure 5). We found no significant LPC effect in HC participants (p (28) = 0.285, p= 0.389, Cohen’s d= 0.053). In contrast, the PD participants showed a significant LPC between 556 to 984ms (p (28) = -2.777, p= 0.005, Cohen’s d= -0.516). As this analysis was exploratory and not part of the primary mixed-effects modelling framework, it should be interpreted with caution. The results suggest a potential increase in late-stage control-related activity in PD participants, although this finding is exploratory.

**Figure 5.**
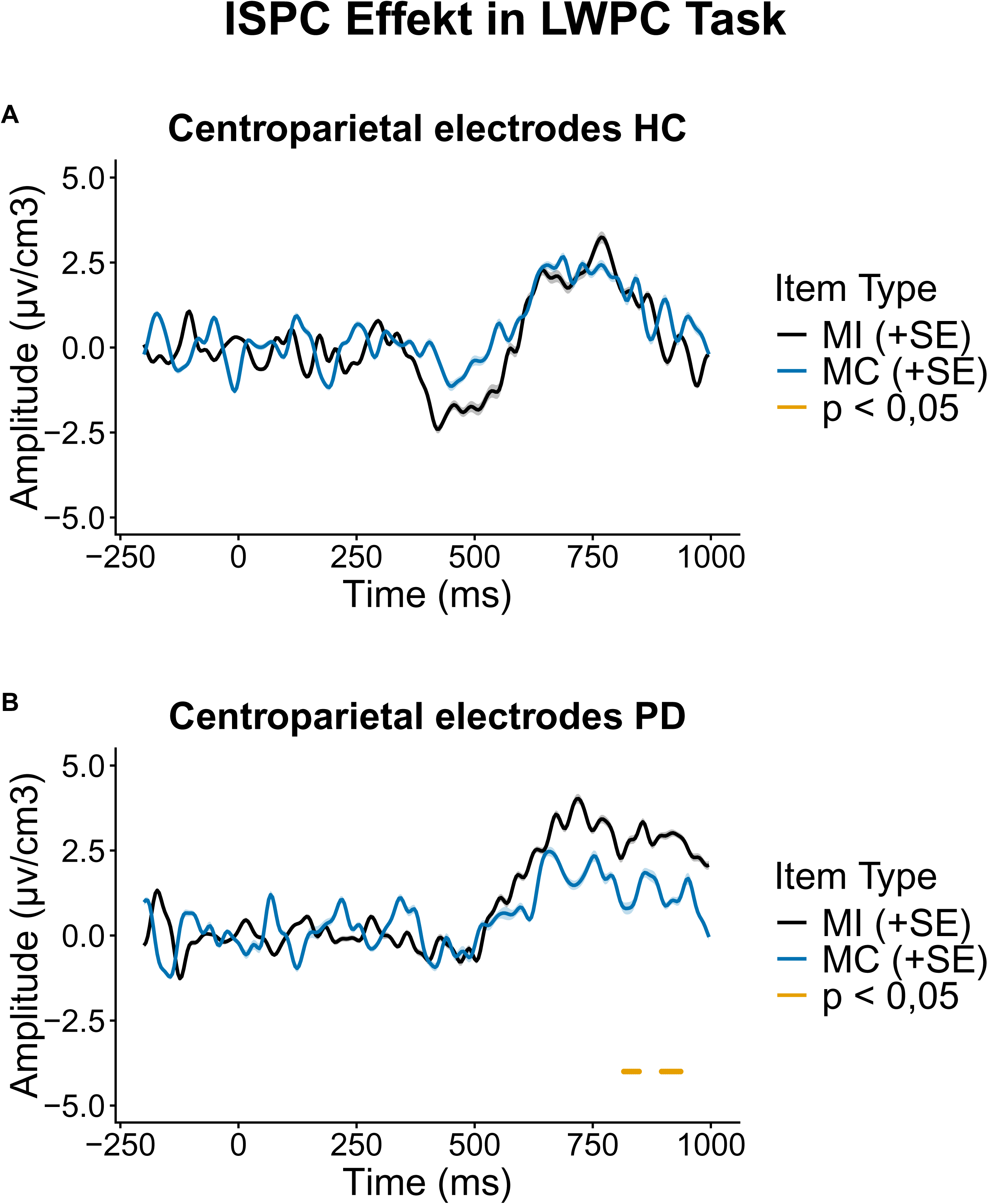
ERP Results for the ISPC effect in LWPC task: Grand average ERP waveforms recorded from centroparietal channels for Healthy Controls (HC, panel A) and Parkinson’s Disease patients (PD, panel B). The x-axis represents time (ms), 0ms indicates the event onset, and the y-axis shows amplitude (uv/cm3). Black and blue lines correspond to different item types (MI and MC), with shaded areas representing the standard error (SE). Orange horizontal markers indicate time windows with significant differences between conditions (p < 0.05). Statistical analyses were conducted using linear mixed-effects models (LMMs) with Group (HC vs. PD) and Condition (MI vs. MC) as fixed effects and random intercepts for participants. Sample size was N = 28 per group. Each waveform represents the grand-average ERP across participants; thus, values reflect mean amplitudes per time point across individuals.

For the centroparietal N450, the linear mixed-effects model revealed no significant main effect of Group (F(1,84) = 2.93, p = .091), no main effect of Condition (F(1,84) = 0.0015, p = .969), and no Group × Condition interaction (F(1,84) = 0.43, p = .513), indicating no reliable modulation of N450 amplitudes in this region.

## Discussion

This study investigated adaptive control processes in healthy participants and individuals with PD using a numerical Stroop task that included proactive and reactive control manipulations. To minimize bias from lower-level learning processes, we distinguished between manipulated inducing items and unbiased diagnostic items. This distinction allowed us to ensure that the behavioural performance in adaptive control was not confounded by the learning effects of inducer trials. We found significant differences in signatures of adaptive control processes between PD and HC. Specifically, our analyses revealed a marked impairment in proactive control in PD, as evidenced by ERPs. Importantly, the ERP findings suggest preserved reactive control-related processes in PD. The neurophysiological data suggest that deficits in proactive control were compensated for by increased reliance on reactive control.^19,27^

In this study, ERP correlates were evaluated in the context of proactive and reactive control in people with PD. Previous research, such as Rustamov et al.,^27^ used EEG to examine the congruency sequence effect (CSE), which primarily reflects reactive control. However, the CSE inherently conflates reactive and proactive processes because improved performance following incongruent trials may result from increased attentional adjustments (reactive control) or from sustained conflict expectation (proactive control). As a result, it is difficult to disentangle whether observed adaptations are triggered by the previous trial (reactive) or reflect anticipation and preparation for upcoming conflict (proactive).^41^ In contrast, the LWPC and ISPC effects provide a more precise differentiation because they directly measure proactive and reactive control. Our study offers a more nuanced characterization of cognitive control processes by distinguishing between diagnostic and inducer trials. This provides an advantage compared to the use of the CSE as a pure measure of reactive control, because in the CSE the effects can be influenced by various mechanisms, including proactive strategies, item-specific learning processes and episodic memory.^3,42^

Importantly, our ERP findings extend previous EEG research in PD by demonstrating that proactive and reactive control processes can be dissociated at the neurophysiological level. While Rustamov et al.^27^ reported preserved conflict adaptation effects in PD, the present findings indicate that such preserved adaptation may primarily reflect intact reactive control mechanisms rather than preserved proactive control. This interpretation is consistent with theoretical accounts proposing that PD is characterized by impaired maintenance of goal-relevant contextual information due to dysfunction within frontostriatal dopaminergic circuits, whereas stimulus-driven adjustments remain relatively preserved.^6,8,43^

### Proactive control in PD participants

Behavioral data indicated that both PD and HC participants exhibited proactive control, as reflected in significant LWPC effects for both inducer and diagnostic items. The presence of LWPC effects in diagnostic items suggests that control adjustments generalized beyond the items that induced the conflict manipulation and therefore reflect context-dependent control rather than simple stimulus-response learning.^2,6,15^ Although LWPC effects were numerically larger in HC participants, the analyses did not provide evidence for a significant group difference in conflict adaptation. Overall, these findings suggest that the ability to adapt cognitive control to changing conflict contexts is largely preserved in PD at the behavioural level.

Importantly, exploratory analyses within the PD group indicated that disease severity modulated proactive control: UPDRS scores were positively associated with the LWPC effect (β = 0.012, p = 0.042), whereas MMST and education showed no associations. The association between UPDRS scores and LWPC magnitude indicates that behavioural indices alone may not fully capture the underlying neural alterations associated with proactive control.

The absence of the N450 component in PD participants provides further evidence for a deficit in proactive control. In healthy controls (HC), the N450 was reliably elicited, reflecting anticipatory conflict monitoring and proactive engagement of control. The N450 has been linked to conflict monitoring processes in medial frontal regions, particularly the anterior cingulate cortex, which signals the need for cognitive control.^42^ This aligns with prior work linking the N450 to early conflict detection and sustained control settings.^34,35^ In contrast, the lack of N450 in PD suggests that patients did not engage in anticipatory control adjustments, even when the task context signalled a high likelihood of conflict. This supports the view that PD is associated with a specific impairment in proactive control mechanisms,^4^ rather than a general deficit in performance. This interpretation is consistent with evidence for altered recruitment of frontostriatal control networks in PD.^43,44^ Taken together, these findings suggest that while healthy controls flexibly adapt their control settings in response to conflict frequency PD participants show a diminished ability to engage proactive control, particularly when generalization beyond the specific stimulus context is required. This pattern aligns with a previous study suggesting that proactive control is selectively impaired in PD.

### Reactive control in PD participants

To disentangle item-specific from context-driven control adjustments, we examined the item-specific proportion congruency (ISPC) effect using both inducer and diagnostic items. While an ISPC effect was observed at the level of inducer items in both groups, no corresponding effect was found for diagnostic items in either HC or PD participants.

A lack of transfer of ISPC effects to diagnostic items was observed in the present dataset. While this pattern could in principle be interpreted as a reduced ability to generalize context-dependent control adjustments, we consider a more parsimonious explanation related to task sensitivity. Importantly, an independent analysis of a young healthy control sample (CY; age range 20–30 years), using the identical experimental paradigm, revealed the same pattern: robust ISPC effects in inducer trials but no reliable transfer to diagnostic items (see Supplementary Table 5). This suggests that the absence of transfer is unlikely to reflect a specific impairment in the Parkinson’s group, but rather a limitation of the diagnostic manipulation itself. Consequently, conclusions regarding reactive control should be restricted to inducer-based measures.

One possible factor contributing to the limited diagnostic transfer may relate to the nature of the stimulus manipulation. The differentiation between numerical magnitudes (e.g., "small" numbers: 1– 4 vs. "large" numbers: 6–9) may not have been sufficiently robust to support stable generalization of control settings across item sets, thereby reducing the sensitivity of the diagnostic condition. While the brain readily differentiates between broad categories such as humans and animals, it may not automatically distinguish between numerical magnitudes (e.g., small vs. large numbers). Future studies should therefore further evaluate the role of numerical magnitude in ISPC paradigms or consider alternative implementations that ensure robust category representation.

The ERP analysis revealed a significant late positive component (LPC) in both the HC and PD groups, reflecting the engagement of reactive neural networks. The LPC has been linked to later stages of conflict processing, including response selection, conflict resolution, and the updating of task-relevant representations following conflict detection.^12,42,45^ Unlike the N450, which is thought to reflect conflict monitoring, the LPC is commonly interpreted as reflecting the implementation of control after conflict has been detected. In addition, reactive control requires less working memory capacity and is therefore easier to manage than proactive control.^34^ The preservation of LPC modulation in PD despite the absence of N450 effects suggests that post-stimulus control processes remain relatively intact. This pattern is consistent with the Dual Mechanisms of Control account, which proposes that proactive and reactive control rely on partly separable neural systems.^6^

### Adaptive control in Event-related potentials

The ERP analysis presented here offers an additional perspective on cognitive control deficits in PD, particularly regarding proactive control. While theta-band activity is a well-established marker of conflict monitoring and cognitive control,^36,46^ ERPs provide the temporal resolution to analyse proactive and reactive control mechanisms. In this study, the absence of a clear N450 in PD participants directly indicates impaired proactive control, which may not have been as evident in theta-band analyses alone. Additionally, ERP components like the N450 and LPC offer more specific insights into different stages of cognitive processing, whether deficits stem from conflict anticipation, resolution, or reactive adjustments.^6,37^ This highlights the value of ERPs in capturing the neural mechanisms underlying cognitive control deficits.

### Compensation for proactive through reactive control

Since Rodriguez-Raeke et al.^14^ hypothesized a compensatory use of brain regions for adaptive control, we asked whether participants with PD could compensate for the lack of proactive control with reactive control. Therefore, we investigated whether there was evidence of reactive control processing in Parkinson’s participants in the block showing proactive control in HC participants.

For this purpose, we examined the ERPs of the MC_LWPC_ and MI_LWPC_ block in the relevant brain regions of reactive control. The results showed a significant difference in the LPC component in centroparietal channels in PD participants. This component was not present in healthy control participants. We hypothesize that this component may indicate compensatory mechanisms, reflecting reactive control processes in participants with PD. The centroparietal distribution of the LPC observed here is broadly consistent with previous work linking this component to frontoparietal networks involved in context updating and adaptive response selection.^21,24^ Given that the LPC is thought to reflect post-conflict evaluation and adaptive response processes, the enhanced LPC activity may indicate increased reliance on reactive control when proactive control mechanisms are compromised. This would also explain why there was an LWPC effect in the behavioural data of participants with PD.

### Explanations for deficits observed in Parkinson’s disease

The observed deficit of proactive control, which likely depends on the sustained goal maintenance in the lateral prefrontal cortex (PFC), may be particularly vulnerable in PD due to its reliance on dopaminergic signalling.^4^ Degeneration of dopaminergic neurons in the substantia nigra disrupts basal ganglia functioning, including the dorsal and ventral striatum, as well as their projections to the thalamus, which plays an important role in stabilizing and relaying goal-relevant information back to the PFC.^47–49^ As a result, PFC-striatal interaction may be weakened, reducing the ability to maintain goal-relevant information over time and to prepare for anticipated conflicts. Reduced dopamine availability may further limit sustained activation, increasing reliance on reactive control, which appears less dopamine-dependent and may be supported by posterior cortical regions and the hyper-direct pathway, a fast route from prefrontal cortex to the subthalamic nucleus that enables rapid, stimulus-driven inhibition.^50^ Neuroanatomically, such reactive adjustments have been associated with interactions between medial frontal, parietal, and subcortical regions involved in conflict resolution and response selection.^42,51^ The preserved LPC modulation observed in the present study may therefore reflect the continued availability of these stimulus-driven control mechanisms despite degeneration of frontostriatal circuits supporting proactive control.

This pattern is consistent with our ERP results, where the absence of N450 but presence of LPC modulation in PD suggests impaired proactive control alongside relatively intact reactive control processes. This interpretation is broadly consistent with previous studies suggesting that cognitive deficits in PD are more pronounced for processes relying on sustained goal maintenance than for stimulus-driven adjustments.^43,44^ Similarly, Rustamov et al.^27^ reported preserved behavioural conflict adaptation in PD despite altered neural processing, which may reflect continued engagement of reactive control mechanisms.

## Limitations

While this study provides insights into cognitive control impairments in PD, a few limitations should be considered. First, variability within the PD group remains a limitation, as disease severity may influence cognitive control and neural responses. This is partly reflected in our exploratory analysis suggesting a potential association between UPDRS scores and proactive control measures. Future studies should further examine disease-stage effects using larger, stratified samples. Second, the influence of medication could not be addressed in the present study, given the on-drug condition during testing, but could play a role. Third, manipulation of the "small" and "large" numbers in the diagnostic items for the reactive control did not consistently produce the predicted behavioural effects. This issue may have reduced the clarity of the expected cognitive control effects in the diagnostic condition. A more robust task manipulation and prior piloting of number scaling effects would help to address this problem in future studies.

## Conclusion

Our findings indicate that Parkinson’s disease is associated with deficits in proactive control, whereas reactive control appears largely preserved. Although this interpretation remains tentative, the absence of the N450 ERP component in PD participants suggests diminished anticipatory engagement, while the observed LPC may reflect a compensatory recruitment of reactive control mechanism. In any case, the study underscores the importance of distinguishing between proactive and reactive control, as well as between diagnostic and inducer items, to accurately characterize adaptive control processes in PD and inform potential interventions aimed at supporting goal-directed behaviour.

## Supporting information

Supplementary Informations

## Data Availability

Due to the inclusion of potentially identifiable participant information and the lack of participant consent for public data sharing, the data cannot be made publicly available. The experiment and code used for the analysis and experiment can be found at: https://osf.io/j39qn.

## Acknowledgements

We are grateful to Martina Bantel for her organizational support and insightful suggestions. We also thank Wolf Pink for his assistance with the measurements.

## Abbreviated summary

ACC: Anterior cingulate cortex
ASR: Artifact Subspace Reconstruction
CSE: Congruency Sequence Effect
DMC: Dual Mechanisms of Control
EEG: Electroencephalography
ERP: Event-related potential
FCz: Frontocentral electrode (EEG channel)
HC: Healthy controls
ICA: Independent Component Analysis
ISPC: Item-specific proportion congruency
LMM: Linear mixed-effects model
LPC: Late positive complex
LWPC: List-wide proportion congruency
MC: Mostly congruent
MI: Mostly incongruent
MMST: Mini-Mental State Examination (MMSE in some conventions; here MMST as used in manuscript)
N450: Negative ERP component around 300–500 ms associated with conflict processing
OSF: Open Science Framework
PD: Parkinson’s disease
PFC: Prefrontal cortex
P3b: Late positive ERP subcomponent related to context updating
RT: Reaction time
SE: Standard error
UPDRS: Unified Parkinson’s Disease Rating Scale

