## Supplementary Informations for "Reactive Compensation for Proactive Deficits in Parkinson’s Disease": Supplementary Information.docx

Supplementary information 1: **Task** design: Rules of Pseudorandomization

The following rules are defined for pseudorandomization: (1) A number combination was presented no more than twice in a row, (2) the left or right answer was correct no more than three times in a row. (3) Congruent or incongruent stimuli were presented no more than four times in succession. (4) Inducer elements were not shown more than four times in a row, (5) diagnostic elements are always shown alone.

Supplementary information 2**: ERP analysis:** **Collapsed Localizer Approach**

We created a grand average difference wave for congruent versus incongruent for the frontal and centroparietal electrodes (CPz, CP1, CP2, Cz, C1, C2, FC1, FC2, FCz), averaged for all conditions and groups using the collapsed localizer approach^1,2^. We then determined the times at which the amplitude of the grand averaged difference curve was 20% of the maximum voltage and used this to define the time window for the N450 component^3^. To determine the time window for the LPC component, we used the same principle but considered the parietal electrodes (P1, P2, Pz, POz, CPz, PPO1h, PPOO2h)^4^.

| Distance | Comparison |
| --- | --- |
| 1 | 1-2, 3-4, 6-7, 8-9 |
| 2 | 1-3, 2-4, 6-8, 7-9 |

**Supplementary Table 1: Overview of the items used in the numerical Stroop task**. The task included number pairs with a numerical distance of 1 or 2, as previously utilized in studies by Dadon and Henik^5^. The items were carefully balanced in terms of numerical representation and overall distribution. For the ISPC manipulation, either pairs of small or large numbers were specifically adjusted.

|  | C_CongruencyHC_ | C_ProportionHC_ | C_InteractionHC_ | C_CongruencyPD_ | C_ProportionPD_ | C_InteractionPD_ |
| --- | --- | --- | --- | --- | --- | --- |
| Incongruent_MI_HC | -0.5 | -0.5 | 0.5 | 0 | 0 | 0 |
| Incongruent_MC_HC | -0.5 | 0.5 | -0.5 | 0 | 0 | 0 |
| Congruent_MC_HC | 0.5 | 0.5 | 0.5 | 0 | 0 | 0 |
| Congruent_MI_HC | 0.5 | -0.5 | -0.5 | 0 | 0 | 0 |
| Incongruent_MI_PD | 0 | 0 | 0 | -0.5 | -0.5 | 0.5 |
| Incongruent_MC_PD | 0 | 0 | 0 | -0.5 | 0.5 | -0.5 |
| Congruent_MC_PD | 0 | 0 | 0 | 0.5 | 0.5 | 0.5 |
| Congruent_MI_PD | 0 | 0 | 0 | 0.5 | -0.5 | -0.5 |

Supplementary **Table 2: Contrast Matrix**

| **Group** | **Control** | **Congruency** | **ItemType** | **N** | **Mean_RT** | **SD_RT** |
| --- | --- | --- | --- | --- | --- | --- |
| HC | ISPC | congruent | Diagnostic | 2387 | 648.7 | 193.0 |
| HC | ISPC | incongruent | Diagnostic | 2312 | 705.4 | 185.7 |
| HC | LWPC | congruent | Diagnostic | 2376 | 650.7 | 196.8 |
| HC | LWPC | incongruent | Diagnostic | 2300 | 716.6 | 210.6 |
| PD | ISPC | congruent | Diagnostic | 2367 | 708.2 | 221.1 |
| PD | ISPC | incongruent | Diagnostic | 2274 | 774.3 | 225.7 |
| PD | LWPC | congruent | Diagnostic | 2371 | 719.5 | 214.7 |
| PD | LWPC | incongruent | Diagnostic | 2259 | 778.7 | 221.1 |
| HC | ISPC | congruent | Inducer | 5628 | 624.0 | 175.7 |
| HC | ISPC | incongruent | Inducer | 5535 | 683.4 | 182.2 |
| HC | LWPC | congruent | Inducer | 5617 | 645.9 | 204.6 |
| HC | LWPC | incongruent | Inducer | 5515 | 704.2 | 203.9 |
| PD | ISPC | congruent | Inducer | 5562 | 707.1 | 221.9 |
| PD | ISPC | incongruent | Inducer | 5431 | 761.7 | 228.7 |
| PD | LWPC | congruent | Inducer | 5581 | 696.9 | 210.2 |
| PD | LWPC | incongruent | Inducer | 5442 | 756.1 | 213.8 |

**Supplementary** Table 3. Descriptive reaction times (ms) and standard deviations. Mean reaction times (RTs) and standard deviations (SDs) are shown for healthy control participants (HC) and participants with Parkinson’s disease (PD) as a function of proportion congruency condition (LWPC, ISPC), congruency (congruent, incongruent), and item type (inducer, diagnostic). Values are based on correct trials with RTs > 200 ms after exclusion of practice trials.

|  | **b** | **SE** | **t** | **p** | **r** | **b** |
| --- | --- | --- | --- | --- | --- | --- |
| Group (PD vs. CO) | 0.106 | 0.047 | 58.6 | 2.24 | **0.029** | 0.281 |
| Congruency | 0.110 | 0.005 | 64353.1 | 22.62 | **< .001** | 0.089 |
| Analysis type (main_incon) | 0.005 | 0.005 | 64353.7 | 1.09 | 0.276 | 0.004 |
| Analysis type (MC) | 0.009 | 0.004 | 64353.6 | 2.45 | **0.014** | 0.010 |
| Analysis type (MI) | 0.029 | 0.005 | 64353.2 | 6.01 | **< .001** | 0.024 |
| Group × Congruency | -0.016 | 0.007 | 64353.1 | -2.29 | **0.022** | 0.009 |
| Group × Analysis type (main_incon) | 0.021 | 0.007 | 64353.6 | 3.01 | **0.003** | 0.012 |
| Group × Analysis type (MC) | -0.011 | 0.005 | 64353.8 | -1.99 | **0.046** | 0.008 |
| Group × Analysis type (MI) | -0.014 | 0.007 | 64353.2 | -2.10 | **0.035** | 0.008 |
| Congruency × Analysis type (main_incon) | -0.048 | 0.007 | 64353.1 | -7.01 | **< .001** | 0.028 |
| Congruency × Analysis type (MC) | 0.007 | 0.007 | 64353.1 | 1.06 | 0.291 | 0.004 |
| Congruency × Analysis type (MI) | -0.045 | 0.007 | 64353.0 | -6.60 | **< .001** | 0.026 |
| Group × Congruency × Analysis type (main_incon) | 0.012 | 0.010 | 64353.1 | 1.28 | 0.199 | 0.005 |
| Group × Congruency × Analysis type (MC) | -0.009 | 0.010 | 64353.1 | -0.92 | 0.356 | 0.004 |
| Group × Congruency × Analysis type (MI) | 0.005 | 0.010 | 64353.0 | 0.52 | 0.606 | 0.002 |

**Supplementary Table 4:** **Full fixed-effects parameter** estimates for the linear mixed-effects model examining the effects of group, congruency, and analysis type on log-transformed reaction times. Estimates (b), standard errors (SE), degrees of freedom (df), t-values, p-values, and effect sizes (r) are reported. Random intercepts were specified for subjects and items. The main manuscript reports only the theoretically relevant effects; the present table contains the complete model output for transparency and reproducibility.

| **Model** | **Condition** | **Effect type** | **Estimate** | **SE** | **z** | **p** |
| --- | --- | --- | --- | --- | --- | --- |
| LWPC | Inducer | Interaction | -0.054 | 0.008 | -6.68 | <.001 |
| LWPC | Diagnostic | Interaction | -0.043 | 0.010 | -4.25 | <.001 |
| ISPC | Inducer | Interaction | -0.031 | 0.008 | -3.75 | <.001 |
| ISPC | Diagnostic | Interaction | -0.003 | 0.010 | -0.33 | .745 |

**Supplementary Table 5:** **Exploratory analysis of ISPC and LWPC effects in an independent young healthy control sample (CY; age range 20–30 years) using identical mixed-effects models.** Robust congruency-by-context interactions were observed in inducer trials for both ISPC and LWPC. However, only LWPC showed partial transfer to diagnostic items, whereas ISPC effects did not generalize. This pattern suggests limited sensitivity of the diagnostic manipulation rather than group-specific impairments.

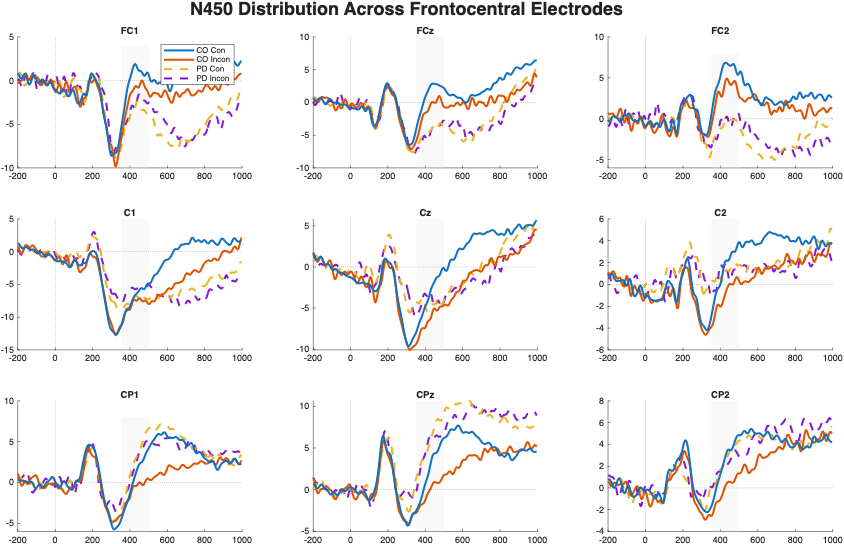

Supplement Figure 1: Grand-average ERP waveforms illustrating the N450 component for congruent and incongruent trials at frontocentral electrode sites (FC1, FCz, FC2, C1, Cz, C2, CP1, CPz, CP2) in healthy controls and Parkinson’s disease participants. The N450 (300–500 ms) shows a more pronounced negative deflection for incongruent compared to congruent trials, with a clear effect in controls and a reduced modulation in the Parkinson’s disease group, indicating attenuated conflict-related processing.

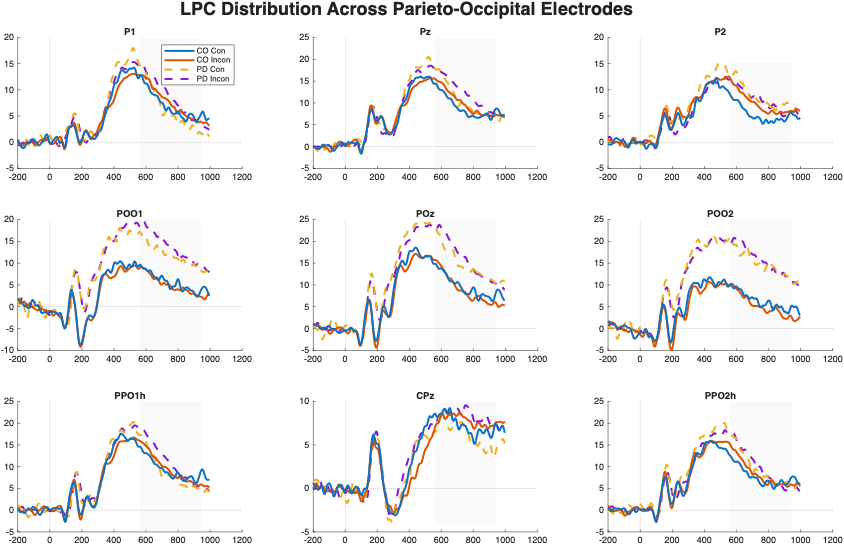

Supplementary Figure 2. Grand-average stimulus-locked ERP waveforms illustrating the Late Positive Complex (LPC) across parieto-occipital electrode sites (P1, Pz, P2, POo1, POz, POo2, PPO1h, CPz, PPO2h). Waveforms are shown separately for controls (CO) and Parkinson’s disease (PD) participants as well as for congruent (Con) and incongruent (Incon) trials. Time is plotted relative to stimulus onset (ms), with negative values indicating the pre-stimulus baseline period.

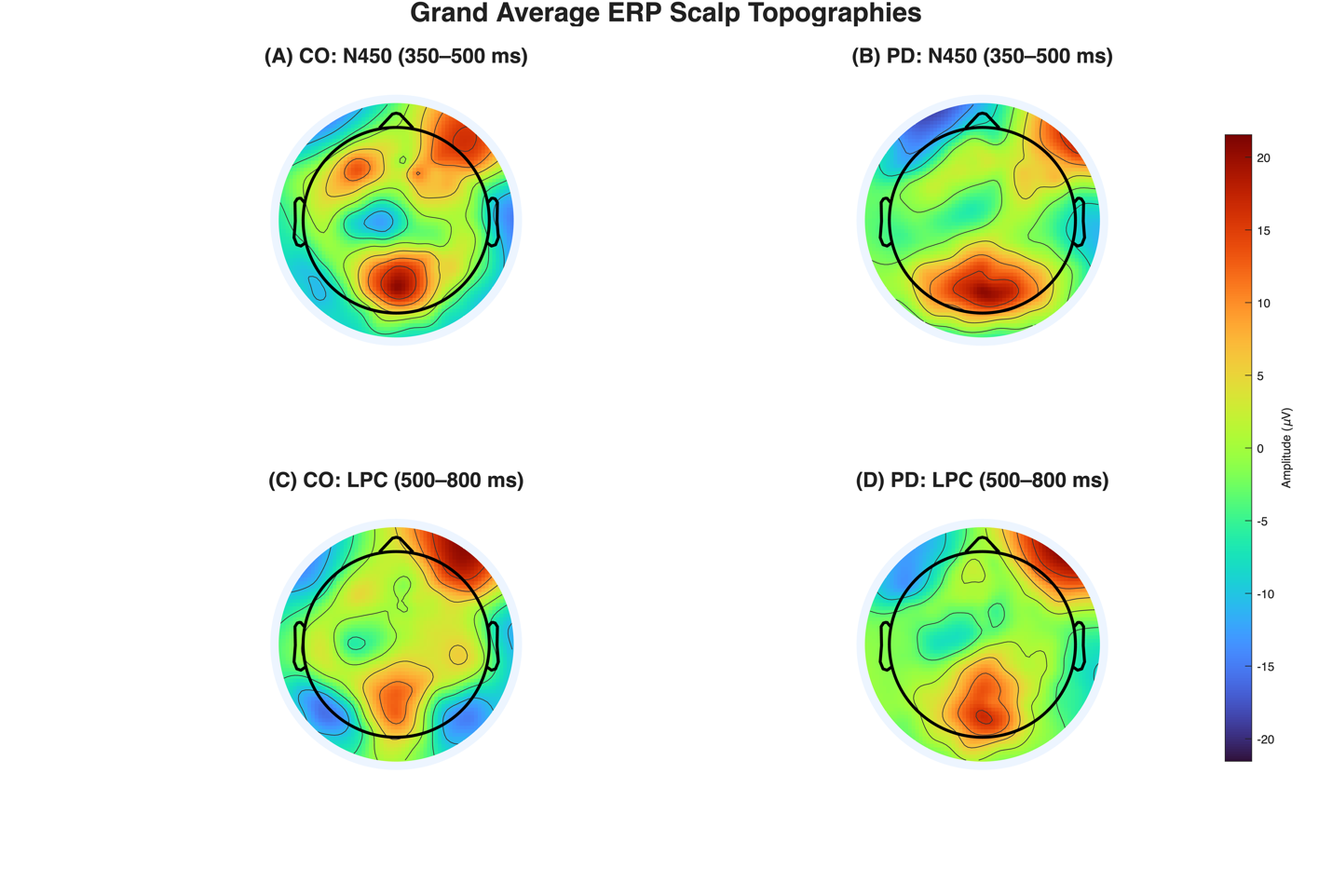

Supplementary Figure 3: Grand-average ERP scalp topographies. Global scalp distributions of the N450 and LPC components for control participants (CO) and Parkinson’s disease (PD) participants. Topographies are shown for the N450 time window (352–500 ms for CO; 350–500 ms for PD) and the LPC time window (560–944 ms) for both groups. The N450 exhibits a frontocentral distribution in both groups, whereas the LPC shows a typical posterior distribution over parieto-occipital regions. All maps are plotted using a common absolute scaling (maximum absolute amplitude across all conditions and components) to allow direct comparison of spatial distributions across groups and components.
